# Behavioural responses to Covid-19 health certification: A rapid review

**DOI:** 10.1101/2021.04.07.21255072

**Authors:** John Drury, Guanlan Mao, Ann John, Atiya Kamal, G James Rubin, Clifford Stott, Tushna Vandrevala, Theresa M Marteau

**Affiliations:** University of Sussex, School of Psychology, UK; Swansea University, Population Data Science, UK; Birmingham City University, Department of Psychology, UK; King’s College London, Department of Psychological Medicine, UK; Keele University, School of Psychology, UK; Kingston University, Department of Psychology, UK; Behaviour and Health Research Unit, University of Cambridge, UK

**Keywords:** Covid-19, SARS-CoV-2, Covid-status certification, health certification, vaccine passport, mandatory vaccination, vaccination certificate, immunity certificate, immunity passport, health passport

## Abstract

**Background:** Covid-status certification – certificates for those who test negative for the SARS-CoV-2 virus, test positive for antibodies, or who have been vaccinated against SARS-CoV-2 – has been proposed to enable safer access to a range of activities. Realising these benefits will depend in part upon the behavioural and social impacts of certification. The aim of this rapid review was to describe public attitudes towards certification, and its possible impact on uptake of testing and vaccination, protective behaviours, and crime.

**Method:** A search was undertaken in peer-reviewed databases, pre-print databases, and the grey literature, from 2000 to December 2020. Studies were included if they measured attitudes towards or behavioural consequences of health certificates based on one of three indices of Covid-19 status: test-negative result for current infectiousness, test-positive for antibodies conferring natural immunity, or vaccination(s) conferring immunity.

**Results:** Thirty-three papers met the inclusion criteria, only three of which were rated as low risk of bias. Public attitudes were generally favourable towards the use of immunity certificates for international travel, but unfavourable towards their use for access to work and other activities. A significant minority was strongly opposed to the use of certificates of immunity for any purpose. The limited evidence suggested that intention to get vaccinated varied with the activity enabled by certification or vaccination (e.g., international travel). Where vaccination is seen as compulsory this could lead to unwillingness to accept a subsequent vaccination. There was some evidence that restricting access to settings and activities to those with antibody test certificates may lead to deliberate exposure to infection in a minority. Behaviours that reduce transmission may decrease upon health certificates based on any of the three indices of Covid-19 status, including physical distancing and handwashing.

**Conclusions:** The limited evidence suggests that health certification in relation to COVID-19 – outside of the context of international travel – has the potential for harm as well as benefit. Realising the benefits while minimising the harms will require real-time evaluations allowing modifications to maximise the potential contribution of certification to enable safer access to a range of activities.

## Background

The current global pandemic caused by SARS-CoV-2 has resulted in wide ranging health, social and economic impacts, including many restrictions on daily movements, contacts, and activities. As testing and immunisation programmes are rolled out, one way of enabling increased access to a wide range of activities is certification of health status. This refers to the action or process of providing an official document – on paper, electronically or other approved medium – indicating that the holder is at low risk of acquiring or transmitting SARS-CoV-2. This could be due to a test-negative result for current infectiousness, a positive antibody test result conferring natural immunity, or vaccination(s) conferring immunity.

Health certification could have many benefits, through enabling greater and safer access to international travel, music, theatre and sports events, and to pubs, restaurants, hotels, and gyms. Allowing people to return to work, meet socially, and fulfil care obligations brings many social, emotional and economic benefits. Indeed, it might be considered unethical to restrict the movements of those who pose minimal risk to others [1, 2]. Depending on how it is applied, health certification could also encourage vaccination uptake [3]. It also has the potential for harm. One concern from a behavioural perspective is that certification may foster an erroneous sense of *no risk* – both in those with and those without certificates – resulting in behaviours that increase risk of infection or transmission. In addition, immunity certification based on a test-positive result for antibodies could have a paradoxical effect on health protective behaviours whereby people deliberately seek infection in order to acquire a certificate [4, 5, 6]. Vaccination certificates could also increase opposition to vaccination in some groups [3]. Concerns have also been raised from ethical and legal perspectives. These include privacy [5], the removal of civil liberties [1, 2], loss of social cohesion by the creation of a new hierarchy [1, 6], discrimination against some social groups [5, 4, 6], and crime, including forgery, cheating, or obtaining documentation or data illicitly [4, 5].

The use of health certificates – also referred to as ‘health’ or ‘vaccine passports’ – is not new. Printed health passes were used in Europe from the late 15th century to allow travel and trade while controlling the spread of plague [7]. They certified only that the bearer had come from a city that was free from plague [8]. The Vaccination Act of 1853 made smallpox vaccination compulsory in Britain for infants. Parents were given a blank certificate of vaccination when registering their child’s birth, to be returned, signed, within three months. Failure to do so resulted in fines and imprisonment [9].

In relation to the current Covid-19 pandemic, certification has been used in China in the form of QR codes allowing entry into public spaces and a range of settings including workplaces, public transport, schools, airports, restaurants and grocery stores [10]. These codes amass data including exposure to places and people at higher risk of transmission. Certification was also used in Slovakia as part of population mass testing for infection. Those testing negative were given a paper certificate and released from strict curfew, thereby allowing return to all workplaces and visits to non-essential shops and restaurants [11, 12]. In the UK, Covid-19 health certification is being planned or being used in limited number of areas, including visits to care homes [13, 14], attendance at football games [15], and some music venues [16]. At the time of writing, Israel is operating a ‘green pass’ scheme in the form of an app which shows whether people have been fully inoculated or have already had the virus [17]. This allows access to gyms, hotels, theatres, and concerts.

The main area where certification (for antigen testing) has been in active use is international travel. The EU has recently announced a ‘digital green certificate’ scheme, enabling those vaccinated, having a recent negative antigen test, or recovered from Covid-19 to travel freely and without quarantine between states within the bloc [18]. The International Air Transport Association has also been developing a digital health pass to “manage and verify the secure flow of necessary testing or vaccine information among governments, airlines, laboratories and travellers” [19]. A number of airlines are using digital health passports, mostly on a trial basis, including British Airways, Virgin Atlantic, and American Airlines [20].

Realising the benefits of health certification in the case of Covid-19 will depend in part upon understanding the possible behavioural and social impacts as a basis for designing systems that mitigate their potential harms. This paper describes the results of a rapid review to examine evidence for such impacts in four areas: (1) public acceptability, (2) effects on uptake of tests and vaccination, (3) impact on behaviours that affect transmission, and (4) crime.

## Methods

A rapid review of the literature was undertaken in accordance with PRISMA criteria [21] to identify the potential impact of enabling access to activities through certificating for one of three outcomes in relation to covid-19 status: (a) negative test results for the virus; (b) positive results on a test indicating immunity; (c) vaccination against Covid-19.

### Search strategy

The search strategy was applied to four peer-reviewed databases -- Web of Science (Core Collection, BIOSIS Citation Index, BIOSIS Previews, KCI-Korean Journal Database, Medline, Russian Science Citation Index, SciELO Citation Index), Ovid (Journals@Ovid, Global Health), Scopus, and APA PsycINFO -- and four pre-print databases -- SocArXiv, MedRXiv, PsyRXiv, and SSRN. These databases were selected based on their coverage of public health topics.

For the grey literature, a search was conducted through the websites of public polling companies such as YouGov and Ipsos MORI; websites detailing public, private and third-sector research projects into Covid-19; and academic websites. Many of these websites were initially identified through a web search using Google Advanced. References and forward citations of relevant articles were also searched.

The search used terms related to the following keywords: “Vaccination certificate”, “Test to enable”, “Immunity certificate”, “Immunity passport”, “Health passport”, “Health certificate”, “Health pass”, “Digital health pass”, “Health code”, “Health code app”, “Immunity-based license”, “Risk-free certificate”, “Mandatory vaccination”, “Mandatory immunisation”, “Compulsory vaccination”. Searches of peer-reviewed databases were conducted on 24^th^ November 2020. All other searches were conducted on a continual basis between the 24^th^ November 2020 and 28^th^ of December, 2020.

### Inclusion and exclusion criteria

The following inclusion criteria were used:

i. Participants: Studies were included if they investigated either attitudes towards health certification, or the behavioural consequences of introducing health certification, in relation to COVID-19 and other infectious diseases. Studies were excluded if they concerned health certification for children^1^ or healthcare workers.
ii. Interventions: The action or process of providing an official document, or “certificate”, which grants access to activities based on (a) negative test results for infectious disease (b) positive immunity test results (c) vaccination against infectious disease. We also included studies of public views of mandatory vaccination given that mandation can only be enforced with some kind of check – i.e. certification.
iii. Comparisons: Certification (for different activities) vs no certificate given.
iv. Outcomes: Beliefs and attitudes towards health certification; behavioural and social outcomes of certification.
v. Study Design: No exclusions were made based on study design.
vi. Characteristics: Studies were included if they presented novel data and were published between January 2000 and the present day.

Given the relative paucity of evidence, we took a liberal approach to the inclusion criteria, which allowed us to add a small number of studies judged to be relevant that were not identified in the search (e.g., a study on the phrasing of test results).

### Risk of bias

Risk of bias was measured using the Mixed Methods Appraisal Tool (MMAT http://mixedmethodsappraisaltoolpublic.pbworks.com; see 22) evaluating studies on five dimensions based on the study method. Studies were rated as good quality if they scored four or more out of five; moderate quality if they scored three out of five; and poor quality if they scored two or less out of five. See Supplementary Information for details: https://osf.io/357kt/?view_only=475cd0776a274e6bbc74f95e1eecd0e0

## Results

### Search Results

The search of peer-reviewed databases identified 6292 citations; searches of pre-print databases identified a further 18 citations. Of these, 1133 were duplicates and were removed, with 5178 citations remaining. A search of the grey literature identified 25 additional citations. After title, abstract and full-text screening of all citations, 33 were judged to meet the eligibility criteria. Additionally, 1 article was identified through backward referencing (see Figure 1).

**Figure 1.**
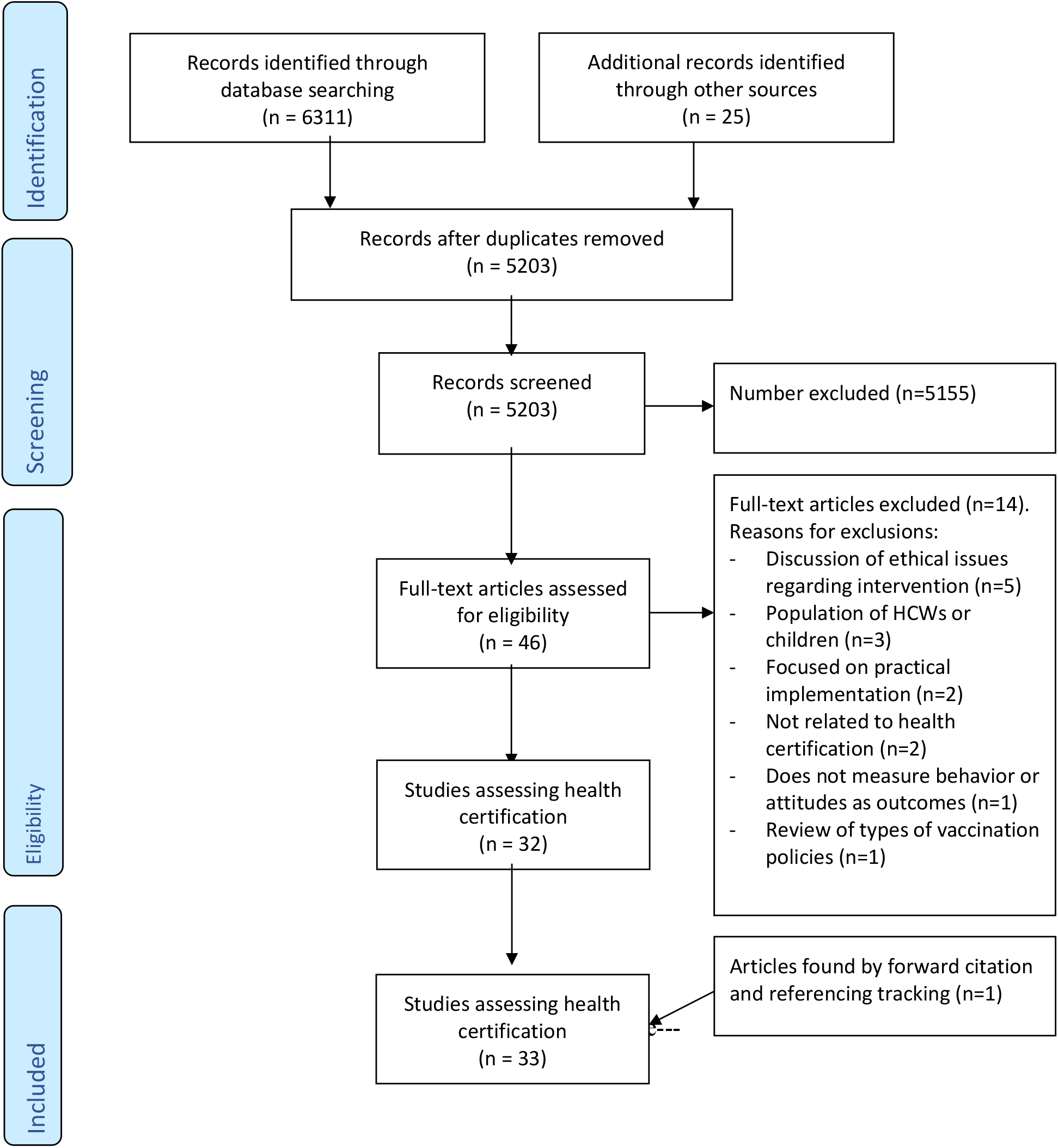
PRISMA Flowchart depicting the selection of studies for the systematic review.

32 of the 33 studies used quantitative methods, with one study using qualitative (narrative) methods. Of the quantitative studies, the majority were cross-sectional surveys (n=30), with the remaining being experimental in design (n=3). Studies were conducted in a variety of countries: Germany (9), UK (10), US (3), Australia (2), Canada (1), Nigeria (1), Poland (1), Romania (1), Spain (1) and Switzerland (1). Three studies drew large samples from several countries (15, 19 and 11 respectively). Of the types of intervention that were the focus of studies, 2 concerned test-negative result for current infectiousness, 14 concerned test-positive for antibodies conferring natural immunity, and 17 concerned vaccination(s) conferring immunity. The majority of studies related to Covid-19 (31), with one concerning yellow fever, and one other concerning flu vaccinations.

### Risk of Bias analysis

Using the MMAT, the mean average risk of bias score was 1.5 from a maximum of 5 (where a higher score means lower risk of bias). In many cases authors did not describe studies in sufficient detail for an evaluation to be made (see Supplementary Information: https://osf.io/357kt/?view_only=475cd0776a274e6bbc74f95e1eecd0e0). Based on the available information, 15 of the studies were rated as low quality, 14 as medium, and three as high.

### Overview

We present a narrative analysis of the results on the impacts of certification in four areas: (1) public acceptability; (2) effects on uptake of tests and vaccination; (3) impact on behaviours that affect transmission and (4) crime. All results are summarized in Table 1.

**Table 1:** Study characteristics.

| **Author**<br>**(Date)**<br>**Country of Study** | **Study Design** | **Participants** | **Data collection**<br>**period** | **Disease** | **Intervention** | **Findings** |
| --- | --- | --- | --- | --- | --- | --- |
| Adepoju, P.<br>(2019)<br>Nigeria | Narrative | N/A | N/A | Yellow<br>fever | Vaccination<br>certificate | • Yellow fever is the only disease specified by WHO for which countries can require proof of vaccination from travellers.<br>• The shortage of vaccines in Nigeria, combined with yellow fever epidemics, has led to the creation of a black market for counterfeit vaccination cards. |
| Behavioural Insights<br>Team (2020)<br>UK | Experiment | 4765 | 13/11/2020 -<br>16/11/2020 | Covid-19 | Covid Test | • A negative personal test result for COVID-19 decreases stated intention to comply with government guidance by 2ppt. Accompanying negative results with a certificate decreases stated intention to comply by a further 5ppt.<br>• A negative test result decreases the proportion of participants saying they would not meet friends by 7ppt. Accompanying negative results with a certificate further decreases this by 6ppt. |
| Betsch, C., et al<br>(2020a)<br>Germany | Experiment | 993 | 23/06/2020 -<br>24/06/2020 | Covid-19 | Mandatory<br>vaccination | • A hypothetical compulsory vaccination against Covid-19 had a negative effect on the willingness to undertake a voluntary vaccination against influenza.<br>• Compulsory vaccination against Covid-19 (compared to voluntary vaccination) led to greater irritation, especially a) amongst participants who had an attitude that vaccinations should be voluntary and b) if the importance of high vaccination rates were not communicated.<br>• Irritation then had a negative effect on willingness to accept the flu vaccination. |
| Betsch, C., et al<br>(2020b)<br>Germany | Survey | 1007 | 05/05/2020 -<br>06/05/2020 | Covid-19 | Immunity<br>certificate | • 48.6% of respondents disagreed with the introduction of an “immunity card”, with around 25.6% agreeing.<br>• 67% felt that those with immunity cards should have no privileges; 13% thought they should have freedom of movement; 8% fewer restrictions; 6% removal of the mask requirement.<br>• Further analyses showed that the respondents would not intentionally get infected in order to receive an immunity pass (no data shown to confirm this) |
| Betsch, C., et al<br>(2020c)<br>Germany | Survey | 1014 | 12/05/2020 -<br>13/05/2020 | Covid-19 | Immunity<br>certificate | • 45.1% of respondents disagreed with the introduction of an "immunity card", with 26.2% agreeing. |
| Betsch, C., et al (2020d)<br>Germany | Survey | 972 | 19/05/2020 - 20/05/2020 | Covid-19 | Immunity certificate | • 45.2% of respondents disagreed with the introduction of an "immunity card". |
| Betsch, C., et al (2020e)<br>Germany | Survey | 925 | 25/05/2020 - 26/05/2020 | Covid-19 | Immunity certificate | • 45.9% of respondents disagreed with the introduction of an "immunity card". |
| Betsch, C. & Bohm, R. (2016)<br>Germany | Experiment | 297 | Not known | Not specific | Mandatory vaccination | • Compulsory vaccination increased the level of anger among individuals with a negative vaccination attitude, whereas voluntary vaccination did not. This led to a decrease in vaccination uptake by 39% in the second voluntary vaccination (reactance).<br>• Making selected vaccinations compulsory can have detrimental effects by decreasing the uptake of voluntary vaccinations |
| Bricker, D (6 Nov, 2020)<br>Canada | Survey | 1,000 | 23/10/2020 - 26/10/2020 | Covid-19 | Mandatory vaccination | • Support for mandatory vaccinations has fallen from 72% in July to 61% in October. |
| COSMO (2020)<br>Germany | Continuous surveys approx. each fortnight | Varied: around 1000 each time. | 14/04/2020 - 15/12/2020 | Covid-19 | Mandatory vaccination | • Vaccination intent has gone from 79% on 14/04/2020 to 49% on 15/12/2020.<br>• Support for mandatory vaccination has gone from 73% on 14/04/2020 to 36% on 15/12/2020. |
| Dennis, S. et al (2020)<br>Australia | Survey | 1169 | 15/04/2020 | Covid-19 | Immunity certificate | • Final support for immunity passports: 10.6% not at all, 49.9% slightly to moderately, 25.1% a lot to fully.<br>• Likelihood of self-infection: 70.4% not at all, 21.7% slightly to moderately, 7.8% a lot to extremely. |
| Feleszko, W. et al (2020)<br>Poland | Survey | 1066 | 02/06/2020 - 09/06/2020 | Covid-19 | Vaccination certificate | • Respondents indicating that they do not plan to vaccinate if the COVID-19 vaccine becomes available (N=301) were confronted with a list of eight different hypothetical reasons to vaccinate. When asked if any of the reasons would sway them to be in favor of being vaccinated, the majority (51%) answered that none of the presented reasons would change their decision. The list of presented reasons included both "High penalties for not vaccinating myself or my child (e.g. 5000 PLN equivalent ca. 1000€)" and "It is not possible to enter some countries without a vaccination certificate". |
| Garret, P. et al (2020)<br>Australia | Survey | 449 | 07/05/2020 | Covid-19 | Immunity certificate | • Final support for immunity passports: 10.6% not at all, 49.9% slightly to moderately, 25.1% a lot to fully.<br>• Likelihood of self-infection: 69.7% not at all, 22.6% slightly to moderately, 7.8% a lot to extremely. |
| Graeber, D., et al<br>(2020)<br>Germany | Survey | 851 | 08/06/2020 -<br>04/07/2020 | Covid-19 | Mandatory<br>vaccination | • 70% of respondents would voluntarily be vaccinated against Covid-19.<br>• 51% of interviewees are against and 49% in favour of mandatory vaccination.<br>• The approval rate for mandatory vaccination is significantly higher among those who would get vaccinated voluntarily (59%) than those who would not be (27%).<br>• Willingness to voluntarily be vaccinated is positively correlated with men, age, education, household income.<br>• Mandatory vaccination is rejected with higher probability by women, but favoured by older people. Approval is negatively associated with neuroticism, and positively associated with subjective probability of contracting life-threatening Covid-19. |
| Haney, C. & Laughlin, G.<br>(2020)<br>US | Survey | 1020 | Jun-20 | Covid-19 | Immunity<br>certificate | • 22% of respondents would “probably” or “definitely” seek infection if earning immunity gave access to various opportunities: 14% to go to gatherings greater than 25 people, 13% to visit eldercare facilities, 12% to visit foreign countries, 10% to visit hospital patients, 11% to maintain or access employment at an eldercare facility.<br>• Younger age was significantly positively associated with willingness to seek infection.<br>• 29% of gig workers reported they would seek self-infection to maintain or access employment in eldercare.<br>• 51% of respondents “strongly” or “somewhat” agree that eldercare facilities should be allowed to require immunity certificate from employees. |
| Hearn, A. & Bull, T.<br>(27 Nov, 2020)<br>UK | Survey | 2,000 | Not known | Covid-19 | Mandatory<br>vaccination | • 45% of respondents think the Covid-19 vaccine should be compulsory, with 35% disagreeing entirely.<br>• Of those who did not want to be vaccinated, 19% would do so if they could go to the pub, 35% if they could go on holiday abroad, 28% if they could go to sporting, music or other events.<br>• 71% of people think people arriving in the UK for holiday or business should have a certificate confirming vaccination, 70% think UK residents leaving the country should have a certificate saying they've been vaccinated. |
| IATA<br>(August, 2020)<br>11 countries | Survey | 4700 recent air<br>travellers | Aug-20 | Covid-19 | Covid test | • 88% were willing to undergo a COVID test as part of the travel process, 84% thought it should be required of all travelers. |
| IPSOS Essentials (2020)<br>15 countries | Survey | 14,500 | 27/08/2020 - 30/08/2020 | Covid-19 | Mandatory vaccination | • 39% of respondents in the UK “strongly support” mandatory vaccination; 31% “somewhat support” them.<br>• Support for mandatory vaccinations is generally strongest in countries with the greatest health impact (Brazil, Mexico, India) |
| IRES - Romanian Institute for Evaluation and Strategy (2020)<br>Romania | Survey (Computer Assisted Telephone Interviewing) | 1027 | 13/05/2020 - 14/05/2020 | Covid-19 | Immunity certificate | • Over 4 out of 10 Romanians would be willing to be vaccinated against COVID - 19 once there was an approved vaccine, but 33% say they would not be vaccinated in any form.<br>• 6 out of 10 Romanians would be willing to be tested in exchange for receiving an "immunity passport". |
| Largent, E.A. et al. (2020)<br>USA | Survey | 2730 | 14/09/2020 - 27/09/2020 | Covid-19 | Mandatory vaccination | • 40.9% of respondents found state mandates for adults acceptable, and 44.9% unacceptable.<br>• Slightly more respondents found employer-enforced employee mandates acceptable (47.7% acceptable to 38.1% unacceptable)<br>• Individuals likely to get a COVID-19 vaccine accepted mandates at higher rates than those unlikely to do so (65% vs 17.3% for state-mandated, 72.5% for 22.9% for employer-mandated).<br>• Acceptance of mandate was also positively associated with non-Black respondents and those with a bachelor's degree. No gender differences observed. |
| Lazarus et al. (2020)<br>19 countries | Survey | 13,426 (768 UK) | 16/06/2020 - 20/06/2020 | Covid-19 | Mandatory vaccination | • There is a discrepancy between reported acceptance of a COVID-19 vaccine and acceptance if vaccination was mandated by one's employer: all respondents, regardless of nationality, reported that they would be less likely to accept a COVID-19 vaccine if it were mandated by employers. |
| Lewandowsky, S. (2020)<br>Spain | Survey | 1,500 | 27/04/2020 - 02/05/2020 | Covid-19 | Immunity certificate | • Final support for immunity passports: 17.3% not at all, 60.7% slightly to moderately, 22.1% a lot to fully.<br>• Likelihood of self-infection: 65.6% not at all, 27.3% slightly to moderately, 2.9% a lot to extremely. |
| Lewandowsky, S., et al. (2020)<br>UK | Survey | 1500 | 16/04/2020 | Covid-19 | Immunity certificate | • The majority of respondents did not object to the idea of immunity passports, with over 60% of respondents supporting the idea to varying extents.<br>• Over 60% of respondents wanted an immunity passport for themselves.<br>• Around 20% of respondents considered immunity passports to be unfair and opposed them completely.<br>• 79% of respondents would not consider at all deliberate self-infection to obtain an immunity passport, around 21% considered doing so to varying degrees. |
|  |  |  |  |  |  | • Increased age, greater perceived risk of the disease, greater trust in government were positively associated with acceptance of immunity passports whereas gender had no effect. |
| Lorenz-Spreen, P. et al. (2020)<br>Germany | Survey | 1,109 | 17/04/2020 - 22/04/2020 | Covid-19 | Immunity certificate | Attitudes towards Immunity passports in Germany: Awaiting precise data<br>Available from: https://ai_society.mpib.dev/tracking-app/wave2.html#Immunity_Passports |
| Nehme, M., et al. (2020).<br>Switzerland | Survey | 1425 | 27/05/2020 - 27/06/2020 | Covid-19 | Immunity certificate | • 60% of participants reported that immunity certificates should be offered to the general population.<br>• The contexts where certificates would be perceived as most useful were taking a plane (73%) and entering a country (72%); fewer participants agreed with them being useful for participating in large gatherings (55%) or the right to work (32%).<br>• 55% of participants thought a vaccination should be mandatory and 49% thought a vaccination certificate should be mandatory.<br>• 68% felt there was a potential risk of discrimination.<br>• 28.6% felt there was a risk of deliberate infection to acquire immunity. |
| Qualtrics (Sept 2020)<br>USA | Survey | 1,074 | 21/09/2020 - 24/09/2020 | Covid-19 | Mandatory vaccination | • Requirements that would make respondents "a little more likely" or "a lot more likely" to vaccinate:<br>• To visit a hospital or nursing home: 70%<br>• Travel to another state without quarantining: 70%<br>• Flying: 68%<br>• Going into office to work: 60%<br>• Large gatherings: 59%<br>• Large religious gatherings: 55%<br>• Attend school in person: 51% |
| Redfield & Wilton Strategies (2020)<br>UK | Survey | 1,500 | 16/05/2020 | Covid-19 | Immunity certificate | • 69% of respondents would support a policy of immunity certificates, with 16% against.<br>• 30% of respondents believe an immunity certification policy would implicitly reward those who did not follow social-distancing measures.<br>• 19% of respondents would consider deliberately catching coronavirus in response to a policy of immune certification, whilst 71% would not; 9% were unsure. |
| Savanta:Comres (2020)<br>UK | Survey | 2,090 | 20/11/2020 - 22/11/2020 | Covid-19 | Mandatory vaccination | • Where it is voluntary to receive the vaccine 67% are likely to get it and 23% unlikely. When it is mandatory without legal penalty,<br>less are actually likely to get it (65% to 24%). A legal penalty does not make much difference (65% to 25%). |
| Waller, J., et al. (2020).<br>UK | Survey | 1204 | 28/04/2020 - 01/05/2020 | Covid-19 | Immunity certificate | • Participants did not perceive any difference in risk between the terms Passport, Certificate, or Test for an antibody test.<br>• When using the term Immunity, 19.1% of participants perceived no risk of catching coronavirus compared to 9.8% for the term Antibody.<br>• Perceiving no risk of infection was associated with an intention to wash hands less frequently, but there was no significant associated with intended avoidance of physical contact. |
| YouGov (2 Dec, 2020)<br>UK | Survey | 5351 | 02/12/2020 | Covid-19 | Mandatory vaccination | • 37% of respondents supported government making it legally compulsory for all people in Britain to be vaccinated against Covid-19, with 44% opposing. |
| YouGov (24 Nov, 2020)<br>UK | Survey | 4,311 | 24/11/2020 | Covid-19 | Vaccination certificate | • 72% of people support all airlines instituting a policy of only allowing passengers who can provide proof that they have been vaccinated (42% strongly support, 30% somewhat support). 18% of people disagree and 11% don't know.<br>• Support appears to be correlated with age. No relationship with social grade. |
| YouGov (8 Dec, 2020)<br>UK | Survey | 5396 | 08/12/2020 | Covid-19 | Vaccination certificate | • Those who should have been vaccination should not be subject to any more coronavirus restrictions: 22%<br>• Everyone should be subject to the same coronavirus restrictions until most people have been vaccinated: 66% |
| YouGov/Sky (2 Dec, 2020)<br>UK | Survey | 1706 | 02/12/2020 - 03/12/2020 | Covid-19 | Vaccination certificate | • 50% of respondents would continue to follow coronavirus rules and restrictions just as strictly after having a vaccination; 29% less strictly, 11% not at all.<br>• Opinions of whether it would be "acceptable" to only allow people who have had vaccination to:<br>• Travel by plane: 54% acceptable, 29% not acceptable, 17% unsure<br>• Go to the cinema: 44% acceptable, 37 not acceptable, 20% unsure<br>• Go to a restaurant: 39% acceptable, 43% not acceptable, 19% unsure<br>• Travel on public transport: 36% acceptable, 46% not acceptable, 18% unsure |

Various terms were used to refer to health certification documents, including ‘certificates’, ‘passes’ and ‘passports’, referring to infection, virus, antibodies, immunity and vaccination. The terms used in this section are *infection certification* (based on test-negative results for infection, whether lateral flow test or qPCR) and *immunity certification* (based on either a test-positive result for antibodies or a completed COVID-19 vaccination).

### Public acceptability

Ten studies of public opinion regarding health certification were found. Some asked about access to particular activities while others simply asked about the use of health certification in principle. In addition, eight studies examined attitudes towards mandatory vaccination.

#### Infection Certification

One study surveyed plane passengers (n= 4700) from 11 countries in August 2020. 84% were in favour of infection certification for air travel [24].

#### Immunity Certification: from antibody testing

Four surveys carried out in Germany in May 2020 (ns between 925 and 1014) found that between 45% and 49% disagreed with the introduction of an “immunity pass”, with around 26% agreeing [25, 26, 27, 28]. Two surveys carried out in Australia in April and May 2020 (ns = 1169 and 449) found that ∼11% did not support immunity ‘passports’ or ‘certificates’ at all but ∼75% supported them slightly to fully [29, 30].

Other studies asked about attitudes to immunity certificates for different purposes. Across five studies (n ∼1000 to ∼1700) conducted in four countries between April and December 2020, a majority of participants (54% to 73%) were in favour of the use of immunity certificates, particularly in the context of international travel [31, 32, 33, 34, 35]; a minority (15-20%) strongly opposed their use. One study (n ∼1000) conducted in Germany in May 2020 found the opposite, with more people opposed to than supporting “immunity cards” [36]. A UK survey carried out in December 2020 (n = 1706) reported that while 44% of respondents found vaccination certification acceptable for going to the cinema, this fell to 39% for going to a restaurant [37]. In another UK survey carried out in December 2020 (n = 5396), 22% of respondents said that those who have been vaccinated should not be subject to any more coronavirus restrictions while 68% disagreed [38]. The percentage in favour of immunity certificates for use for the right to work was much lower than in the case of travel. Across three studies in three countries carried out in April – September (n ranging from 1000 to 1500) support ranged from 20% to 51% [31, 33, 39].

There was little information in most studies on how any of the attitudes described above varied across social groups. In the UK, one study found that acceptance increased with age, greater trust in government, and higher perceived risk of COVID-19 [31].

#### Immunity Certification: from vaccination

Only one study of attitudes towards vaccination certificates specifically (n = 4311) was retrieved, conducted in the UK in November 2020, which assessed attitudes towards their use on international flights. 72% supported their use (42% strongly) and 11% strongly opposed them [40]. Support was strongest in older age groups, and unrelated to gender or socioeconomic status.

#### Mandatory vaccination

The terms ‘mandatory’ and ‘compulsory’ vaccination were used in studies to refer to a general requirement by governments for all citizens to be vaccinated, but with the means by which this could be achieved usually left unspecified. A UK survey published in November 2020 (n = 2000) found that 45% of respondents thought the Covid-19 vaccine should be mandatory for everyone, with 35% disagreed entirely [41]. Of those who did not want to be vaccinated, 19% said they would do so if they could go to the pub, 35% if they could go on holiday abroad, and 28% if they could go to sporting, music or other events. A UK survey carried out in December 2020 (n = 5351) also found that 37% supported compulsory vaccination [37]. A survey carried out in Germany in June and July 2020 (n = 851) found that 51% of respondents were against and 49% in favour of mandatory vaccination. The approval rate was significantly higher among those who would get vaccinated voluntarily (59%) than those who would not be (27%) [42]. An American survey carried out in September 2020 (n = 2730) found that acceptance of mandatory vaccination was positively associated with non-Black respondents and those with a bachelor’s degree [43]. An international survey (15 countries) carried out in August found that support for mandatory vaccinations was generally strongest in Brazil, Mexico, and India [44]. A survey in Canada (n = 1000) found that support for mandatory vaccinations fell from 72% in July to 61% in October 2020 [45]. Similarly, a survey in Germany (n = 1169) found that support for mandatory vaccination declined from 73% in April 2020 to 36% in December of the same year [46].

### Uptake of tests and vaccination

Few studies addressed the possible impact of certification on uptake of vaccines or tests. A number suggested that intention to get vaccinated would vary with both the activity enabled by this and the source recommending vaccination.

#### Infection Certification

No studies were found.

#### Immunity Certification: from antibody testing

An online experiment carried out in the UK in April 2020 (n=1204) found that 85% would definitely (56%) or probably (29%) have an antibody test if offered [47].

#### Immunity Certification: from vaccination

One US study (n ∼1000) conducted in September 2020 assessed ‘vaccine rules that would resonate’ [48]. The activities requiring vaccination certification for which most people said they would get a COVID-19 vaccination were: visit a hospital or nursing home (likely uptake rate of 70%), travel to another state (70%), air travel (68%), work (60%), attending large non-religious gatherings (59%), attending large religious gatherings (55%), and attending school (51%). However, a Polish study carried out in June 2020 (n = 1066) [49] found that of those who did not plan to get vaccinated, 51% were not swayed by any reasons. Indirect evidence that certification of vaccination for access to work could reduce uptake of vaccination is provided in a survey of 13,426 adults in 19 countries carried out in June 2020. A baseline of 71% reported that they would be very or somewhat likely to take a COVID-19 vaccine, compared with 61% if the vaccine was recommended by an employer [50]. However, an American survey carried out in September 2020 (n = 2730) found that slightly more respondents found employer-enforced employee mandates acceptable (47.7%) than unacceptable (38.1%) [43]. Those reporting higher levels of trust in information from government sources were more likely to accept a vaccine and take their employer’s advice to do so [50].

#### Mandatory vaccination

Two studies with experimental designs carried out in Germany (ns = 993 and 297) found that if a vaccination were to be presented as compulsory this led to anger (compared to voluntary vaccination) which then had a negative effect on willingness to accept a subsequent vaccine [36, 51]. A UK survey carried out in November 2020 (n = 2090) found that, for mandatory vaccination, the numbers saying they would or would not get a vaccination did not vary depending on legal penalty (65% to ∼25% in each case) [52].

### Impact on behaviours that reduce transmission

The evidence for possible behavioural outcomes of certification is summarised below first, amongst those with a certificate, and second, amongst those without a certificate.

### Those with a certificate

#### Infection Certification

An online experiment (n = 4765) conducted in November 2020 in a UK sample found that intentions to fully follow guidance were 61% for those receiving a negative test result but 56% for those receiving a certificate alongside their negative test result [53]. For those not asked to imagine they had undergone testing, 63% reported fully following guidance.

#### Immunity Certification: from antibody testing

Another UK online experiment (April 2020, n = 1204) assessed the impact of describing a positive test indicating presence of antibodies on risk perception and protective behaviours [47]. Using the term ‘immunity’ as opposed to ‘antibody’ increased the proportion who erroneously perceived they would have no risk of catching coronavirus in the future given an antibody-positive test result, from 9.8% to 19.1%. Perceiving no risk of infection with coronavirus given an antibody-positive test result was associated with an intention to wash hands less frequently.

#### Immunity Certification: from vaccination

A UK survey carried out in December 2020 (n = 1706) found that 50% of respondents said they would continue to follow coronavirus rules and restrictions just as strictly after having a vaccination; 29% less strictly; and 11% not at all [37].

### Those without a certificate

#### Having failed an immunity test

The majority of participants in a Swiss survey said they expected that tests showing an absence of antibodies would encourage people to take more precautionary measures such as wearing of face coverings (76%) and respect for social distance measures (87%) [33].

#### Having not applied for a test

Six studies in four different countries conducted between April and June 2020 (n > 1000 each) reported between 19% [39, 54] and 31% [29, 30, 31, 32] of participants saying that they would likely expose themselves to infection in order to get a certificate. More students compared to other groups reported that they might deliberately infect themselves (58%) [31]. In another study, those who were younger and those who worked in the “gig” economy (29%) were more likely than others to report that they would seek self-infection to maintain or access employment [39]. However, a survey study carried out in Germany in May (n = 1007) found that no respondents reported they would intentionally get infected in order to receive an ‘immunity pass’ (though no data was shown to confirm this) [25]. A further study (in Switzerland) examined expectations of *others’* behaviour and found that 28.6% thought that others might self-infect (respondents were not asked how they themselves might respond) [33].

### Crime

One report [55] described the use of counterfeit certificates for yellow fever. In December 2018, Nigeria and other countries introduced machine-readable yellow fever cards, but cards could still be obtained without evidence of vaccination. More outbreaks were predicted as people continue to carry fake vaccination certificates throughout Africa.

## Discussion

In response to the Covid-19 pandemic, health certification is being used or considered for use to enable increased access to a wide range of activities for leisure, work and travel while minimising risk of transmission of the virus. In part this reflects public attitudes, but it will also shape and be shaped by these attitudes.

Public attitudes were generally favourable towards the use of immunity certificates (based on vaccination or on antibody tests) for international travel, protecting the vulnerable (e.g., in a care home setting), but generally unfavourable towards their use for access to work, educational or religious activities or settings. A significant minority was strongly opposed to certificates of immunity - whether based on antibodies or on vaccination - for any purpose. A minority supported mandatory vaccination. A number of studies suggested that intention to get vaccinated varied with the activity enabled by certification or vaccination (e.g., international travel). There was no evidence in the review that mandatory vaccination including sanction would increase uptake. Some studies suggested that health certification might reduce Covid protective behaviours, including social distancing and handwashing. Making access to settings and activities conditional on antibody test certification may lead to deliberate exposure to infection in a minority, especially among young adults and those in precarious employment. No studies were found suggesting effects of Covid-19 health certification on crime.

This is the first rapid review - to the authors’ knowledge - of studies concerning possible behavioural responses to Covid-19 health certification. Both the quality and quantity of studies was low thus limiting the certainty of any conclusions. The potential benefits of Covid-19 health status certificates – through enabling greater and safer access to international travel and other activities – need to be considered in the context of their potential for harm. At the most general level the evidence reviewed suggests the potential for harms of certification but the nature and scale of these remains uncertain. Also uncertain is how any harms might most effectively be mitigated. The evidence reviewed on the potential impact of certification or mandation on vaccination rates suggests this would not increase vaccination rates and might even reduce them. Mandating vaccinations through various means to reduce or eliminate choice is controversial and much debated particularly in the context of childhood vaccination programmes. While effective in some contexts, other approaches to increasing uptake in children can be as or more effective [56, 57]. Amongst adults, a recent review of vaccination policies found that in 17 of 42 European countries some form of mandation or regulation was used [58].

The limited evidence reviewed here that health certification might reduce Covid-19 protective behaviours is consistent with concerns expressed by WHO that those who believed they had had COVID-19 would reduce their adherence to protective behaviours [59]. It is also consistent with more recent research on behavioural responses to rapid antigen tests and vaccinations against Covid-19. A study of rapid antigen tests in the UK found that around 13% of those receiving a test-negative result reported increasing their interactions with others [60]. Around 40% of those aged over 80 in England reported breaking Covid-19 restrictions in place at the time after receiving their vaccinations [61]. In Israel, the rapid vaccination of much of the adult population was accompanied by a short term rise in Covid-19 infections [62]. These findings are consistent with those vaccinated or certificated as having had the virus reducing their adherence to protective behaviours [63, 64]. Group processes have the potential to amplify these behavioural effects. When those with certificates reduce their protective behaviours, such changes can be seen as normative, leading others in their ingroup – including those without certificates – to do the same [65, 66, 67].

Regardless of the basis for any Covid-19 status certificate issued, certification will indicate that the holder has been deemed to pose a lower risk of infection and perhaps transmission of the virus than those without a certificate. Risks may indeed be lower, but the extent of this is not yet fully known. Importantly a residual risk will remain - i.e., the risk will not be zero. Given relatively low sensitivity of rapid non-PCR tests, those testing negative will have a low but not zero risk of being infectious and transmitting the virus [68]. The extent to which current vaccines prevent infection or reduce transmission of all variants and for how long remains uncertain [87]. Similarly, the degree and duration of protection from infection for those testing positive on current antibody tests is uncertain. [69]

### Maximising benefits and minimising harms

Health certification could enable greater and safer access to a wider range of activities and locations for many people. To realise these benefits while minimising the harms, health certification schemes should be implemented with an evaluation designed in from the outset, and, in keeping with the principles of open science, to include the use of pre-registered protocols. Such schemes should also be designed within a transparent ethical and legal framework to protect privacy, equity and minimise fraud.

Evidence from both testing and vaccination suggests that increased inequalities would be a possible harm of health certification. Participation in NHS Test & Trace is lower in marginalised groups [70, 71] and in areas of high deprivation [72]. The Liverpool mass testing pilot found that uptake in the most deprived areas (16.8%) was half that in the least deprived areas (33.4%) [73]. Data from the UK and other countries suggest that those with lower incomes or education and from minority ethnic groups have lower intentions to undergo COVID-19 vaccination than others [73, 74, 75]. In part these differences in testing and vaccination uptake reflect higher mistrust in government amongst marginalised communities [70, 76, 77, 79]. Stigmatisation, discrimination and racism might also reduce migrants’ and ethnic minority communities’ willingness to come forward [71]. In addition, certification will likely be most readily available as a digital record, which has the potential to exclude those without access to electronic platforms [70]. In summary, disadvantaged groups are underrepresented in those getting tested and vaccinated and would therefore be disproportionately excluded in any covid certification scheme.

Use of the social rewards associated with health certification to encourage take-up of the COVID-19 vaccine [3, 80] might work well with some groups but could backfire with those who are already mistrustful of the authorities. While the authorities in Israel see an incentive-based approach as an alternative to coercion, the scheme has already led to conflict at workplaces [3, 17]. The issue of enforced exclusion of many people from significant areas of social life raises broad issues of justice and fairness and could mobilize a wide constituency. In the 19^th^ century, resistance to the Vaccination Act included violent protests from the working class [9] which contributed to a change in the law allowing exemptions on the basis of conscience [81].

Minimizing the potential harms of certification will require the following. First, there should be equality and equity of access to tests, vaccinations and certificates. Second, there needs to be clear and open communication that is accessible to different communities of the meaning of any results and certificate, the residual risks of infection and transmission, and the implications for individual behaviour. National and local leaders, including community members and community organisations, should be involved in this communication campaign, in line with engagement and public inclusion principles [82, 83]. Finally, practical steps are needed to ensure that no group should be disadvantaged by loss of access to an everyday activity or setting requiring certification, particularly if access to income, health or education will be impacted by these.

### Strengths and limitations of the review

This review included 33 studies pertinent to understanding the possible effects of health certification on public behaviour. To the authors’ knowledge it provides the first overview of studies in this area, with implications for practice and policy.

The review was limited both in scope and quality of studies retrieved. The focus was upon the behaviour of general populations and not upon the behaviour of other relevant actors such as employers or those managing or organising venues and events, entry to which may be dependent upon health certificates. The behaviour of these other actors will also be important in realising benefits of health certification to ensure, for example, that measures designed to reduce transmission at a venue – such as physical distancing – are seen as additional and not substitutes for entrants having a health certificate [84].

Few of the studies included in this view were judged to be high quality. The main reasons for being judged low quality were that it was unclear whether there was a non-response bias; like many surveys conducted during the pandemic, most of the studies featured in this rapid review relied heavily on convenience samples which were not representative [85]. Only three of the studies were peer reviewed at the time of this rapid review. While three were available on pre-print servers, most were unlikely to be published in peer reviewed journals and were often released as public opinion surveys.

All the studies concerning Covid-19 studies relied on self-report measures of behaviour and in response to hypothetical scenarios. This was because these studies were carried out before the introduction of certification.

Most of the studies were from high income countries. Most of the studies did not take process or demographic measures. This restricts what we can conclude about the underlying reasoning behind attitudes such as opposition to covid health certification (e.g., whether privacy concerns vs inequality implications were more important).

Finally, public attitudes and behavioural responses in 2020, when certification schemes were not widely discussed or implemented and populations had less experiences of living with restrictions due to higher prevalence of the virus, will likely change in 2021 as such schemes are introduced or actively considered as an approach to controlling transmission of the virus. (For example, a representative poll carried out in the UK in March 2021 [86] found higher levels of support for vaccine passports for a variety of activities than was found in the 2020 surveys in the present rapid review.

Mindful of these limitations, this review nonetheless provides a starting point for anticipating the potential harms of health certification as a basis for mitigating these to realise the benefits with minimal harms.

## Data Availability

There is no data as this is a rapid review paper. However, a complete list of sources with links can be found here at the OSF site below.

https://osf.io/357kt/?view_only=475cd0776a274e6bbc74f95e1eecd0e0

## Declarations

Ethics approval and consent to participate: N/A

Consent for publication: N/A

Availability of data and materials: N/A

## Authors’ contributions

JD, GM, TM: conception and planning, analysis, writing. GJR, CS, AJ, TV, AK writing.

## Footnotes

1 A recent systematic review of mandatory vaccination for children recently summarized findings as follows: ‘Quantitative studies found little evidence for any factors being consistently associated with support for mandatory vaccination. Qualitative studies found that parents perceived mandatory vaccination schemes as an infringement of their rights and that they preferred universal, compared to targeted, schemes’ [23]

